# Multiplexed Salivary miRNA Quantification for Predicting Severe COVID-19 Symptoms in Children Using Ligation-RPA Amplification Assay

**DOI:** 10.1101/2025.03.10.25323665

**Authors:** Md. Ahasan Ahamed, Zhikun Zhang, Aneesh Kshirsagar, Anthony J. Politza, Usha Sethuraman, Srinivasan Suresh, Steven Hicks, Feng Guo, Weihua Guan

**Affiliations:** Department of Intelligent Systems Engineering, Luddy School of Informatics, Computing, and Engineering, Indiana University, Bloomington, Indiana, 47408, USA; Department of Electrical Engineering, Pennsylvania State University, University Park, PA, 16802, United States; Department of Biomedical Engineering, Pennsylvania State University, University Park, PA, 16802, United States; Division of Emergency Medicine, Department of Pediatrics, Children’s Hospital of Michigan/Central Michigan University, Detroit, MI 48201, United States; Divisions of Health Informatics and Emergency Medicine, Department of Pediatrics, University of Pittsburgh School of Medicine and UPMC Children’s Hospital of Pittsburgh, Pittsburgh, PA 15224, United States; Department of Pediatrics, Division of Academic General Pediatrics, Penn State Neuroscience Institute, Pennsylvania State University, Hershey, PA, 17033, United States

**Keywords:** SARS-CoV-2, salivary miRNAs, ligation-RPA reaction, recombinase polymerase amplification, portable analyzer, portable centrifuge, severe miRNA

## Abstract

Most children with SARS-CoV-2 have mild or asymptomatic symptoms, but some develop severe complications. Early identification of high-risk cases is crucial for timely intervention. Alterations in salivary microRNA (miRNA) levels serve as biomarkers for severity prediction. However, a rapid, non-invasive method is needed to quantify miRNA level changes as an alternative to sequencing. Here, we developed a highly specific and sensitive ligation-recombinase polymerase amplification (RPA) assay for quantifying severe and non-severe miRNAs on a portable platform. The assay begins with a miRNA-templated annealing and ligation-RPA reaction of miR-1273, miR-296, and miR-29. We quantified 100 pM to 1 fM, resolving 1 fM, with 100% specificity. Next, we validated portable extraction against benchtop extraction, achieving R² > 0.85 and r > 0.92 in clinical samples. Finally, testing 154 clinical samples revealed severe miRNA downregulation compared to non-severe cases. The assay achieved high diagnostic accuracy with an AUC of 0.98. This platform enables informed clinical decisions and optimizes resources, especially in resource-limited settings.

## Introduction

The true prevalence of asymptomatic Severe acute respiratory syndrome coronavirus 2 (SARS-CoV-2) infections in children is likely underestimated, as many remain undiagnosed due to mild or absent symptoms. Since the spread of COVID-19 in late 2019 (*1*), the majority of children infected with COVID-19 experience mild or no symptoms. However, a subset develops severe complications (*2*), including multisystem inflammatory syndrome in children (MIS-C) (*3*), severe pneumonia, Acute Respiratory Distress Syndrome (ARDS), cardiac complications, blood clotting disorders, and exacerbation of underlying conditions such as asthma, chronic lung disease, and diabetes. In rare cases, neurological complications such as seizures, encephalopathy, or stroke may also occur. Globally, as of July 12, 2023, SARS-CoV-2 has caused 767.97 million confirmed cases, including about 6.95 million deaths (*4*). According to United Nations Children’s Fund (UNICEF) data, as of December 2023, over 17,400 COVID-19-related deaths were reported among children and adolescents under 20 years of age. Of these, 53% occurred among adolescents aged 10–19, while 47% were among children aged 0–9 (*5*). Severe SARS-CoV-2 presents a significant risk to children’s health and safety (*6*). Although children generally face a lower risk of hospitalization and life-threatening outcomes, the potential for severe disease necessitates the early identification of high-risk cases to enable timely and effective intervention. To improve the outcomes for children with severe SARS-CoV-2 infections, early identification of the infection is imperative for appropriate patient disposition, healthcare resource allocation, and argeted treatment strategies (*7*). Given the need for rapid and non-invasive diagnostic tools, saliva, due to its easy accessibility and non-invasive collection, has attracted considerable attention in developing POCT for various diseases (*8*, *9*). Its widespread acceptance among children and adults makes saliva an excellent choice (*10*). Saliva contains many proteins, DNAs, RNAs, miRNAs, and other components(*11*, *12*). The expression level of miRNA serves as a potential biomarker for diagnosing a wide range of infectious diseases (*3*, *13*, *14*). MiRNAs, a class of short (19-24 bases) endogenous noncoding RNAs (*15*), are pivotal in post-transcriptional gene regulation (*16*). They significantly impact cell signaling and guide the host’s immune response (*3*, *17–19*). The relative abundance of Several salivary miRNAs has been investigated as biomarkers for SARS-CoV-2, which has severe outcomes in children (*3*). Levels of miRNA in saliva could offer distinctive insights into inflammation and the body’s immune response from severe SARS-CoV-2 infection. Consequently, developing multiplexed and quantitative miRNA detection in saliva is critical for the early detection of severe SARS-CoV-2 infection in children. Implementing this through POCT enables effective self-monitoring by patients to manage and control this disease’s spread effectively, especially in environments with constrained resources.

Numerous methods have been established for miRNA detection (*20*, *21*), such as northern bot (*22*), microarray (*23*), reverse transcription-polymerase chain reaction (RT-PCR) (*24*), etc. Northern blot offers high specificity in miRNA analysis, yet it is time-consuming and demands substantial sample quantities. Microarrays excel in large-scale, high-throughput gene expression studies but have higher costs and require complex data analysis (*25–27*). RT-PCR is considered the gold standard for miRNA detection. It is notable for its sensitivity and precision in quantitative detection, necessitating thermal cycling through different temperatures (*9*, *28*). These features make these technologies less feasible for POCT in limited resource areas or for personal use at home. To enable a more straightforward and faster detection of miRNAs, various isothermal nucleic acid amplification methods have been developed, including nucleic acid sequence-based amplification (NASBA) (*29*), strand displacement amplification (SDA) (*30*), rolling circle amplification (RCA) (*31*), as well as the recombinase polymerase amplification (RPA) (*32–35*). Among these methods, RPA has gained widespread attention for its advantages, such as its simplicity, high sensitivity, ability to multiplex with fewer primers, rapid amplification, and operation consistently between 37-42°C (*36*). For miRNA detection, RPA poses challenges for short target length because the recommended length of RPA amplicons is greater than 80 bp (*37*). To solve this issue, PBCV-1 DNA ligase with RPA enables the total length over 80 bp and shows PCR-like sensitivity (*38*). So, miRNA detection using ligation-RPA assay facilitates the rapid and sensitive detection of amplicons in POCT. The main challenge lies in resolving the clinical relevance samples with miRNA concentration near femtomolar (fM) concentration (*39*). Therefore, there is an unmet need for a rapid, non-invasive, and quantitative method to detect miRNA expression level changes in femtomolar level and enable early risk for severe SARS-CoV-2 infection in children.

In this work, we developed a ligation-RPA-based isothermal nucleic acid amplification system for detecting miRNAs associated with severe SARS-CoV-2 infection using a portable analyzer. Our system successfully multiplexed and detected three miRNAs-miR-1273, miR-296, and miR-29 for SARS-CoV-2 infection, quantifying their concentrations to establish clinical relevance. We validated the specificity of the system with both individual and mixed samples, demonstrating that our portable extraction method exhibited performance comparable to benchtop systems. Using this portable platform, we could differentiate between severe and non-severe infections from raw salivary samples, showing the downregulation of specific miRNAs in severe cases. Our platform offers significant potential for early disease diagnosis and practical application in POCT.

## Results

### Design and Validation of Ligation/ RPA Assay

We designed and developed the ligation-RPA assay to enable rapid, sensitive, and specific detection of salivary SARS-CoV-2 miRNAs in children. The assay starts with hybridization between miRNA and probes 1 and 2 (P1 and P2), enhanced by rapid denaturing at 85°C for 2 min, followed by 2 min annealing on an ice bath, as shown in **Fig. 1A**. Following hybridization, P1 and P2 formed a phosphodiester bond between the 3’-hydroxyl(-OH) group of P1 and the 5’-phosphate (PO₄³⁻) of P2, effectively stabilizing the hybridized structure and created 5′-phosphorylated single-strand DNA (ssDNA) (*40*). Next, a ligation PBCV-1 DNA ligase catalyzed and bridged 5′ to 3′ terminals of the complementary sequence of probes P1 and P2 with 3′ to 5′ terminals of the target miRNA oligonucleotides to synthesize a partially double-stranded structure (*38*). The ligation reaction works at 37°C for 10 mins; P1 and P2 were designed to partially hybridize with target miRNA and linked to a long ssDNA (ligation products). The ligation products specifically linked with Forward primer (FP) and Reverse primer (RP) sites in the RPA reaction were effectively and specifically amplified at 37°C for 30 mins. Intercalating dyes SYTO 9 bind to RPA amplicons by inserting themselves between the base pairs of the double-stranded DNA (dsDNA) formed during amplification (*41*). The planar structure of the dye allows it to insert into the hydrophobic space between stacked DNA base pairs, stabilized by hydrophobic and electrostatic interactions with the negatively charged phosphate backbone. Once intercalated, the dye fluoresces, enabling the detection and quantification of the amplified product. miR-1273, miR-296, and miR-29, and P1 and P2 sequences are shown in **Fig. 1B**.

**Fig. 1.**
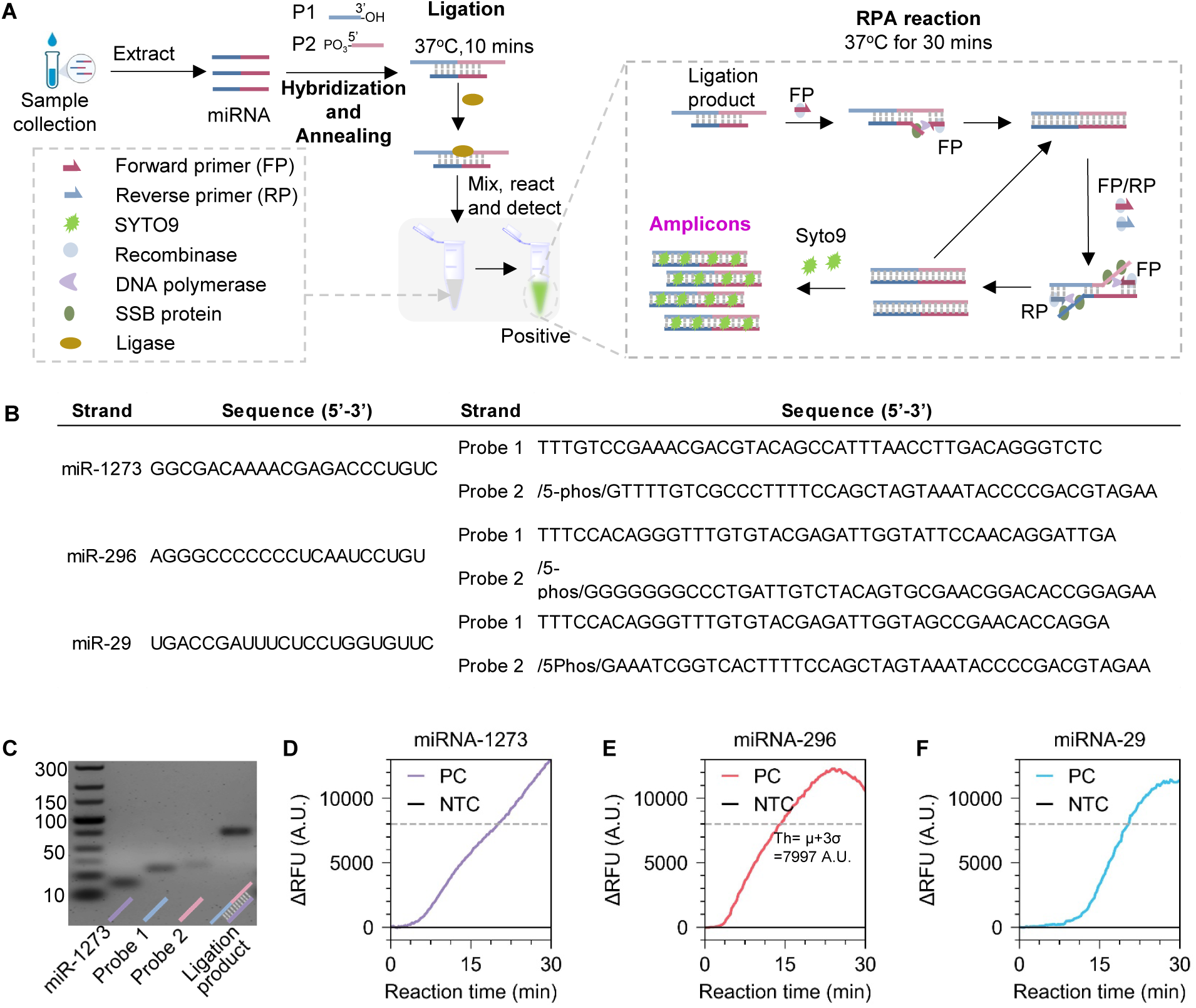
Schematic illustration of ligation-RPA assay for SARS-CoV-2 miR-1273, miR-296, and miR-29 and validation using the benchtop instrument. (**A**) The operation workflow of the ligation-RPA assay. The sample was extracted from saliva, followed by hybridization at 85 °C for 2 min, annealing, ligation reaction at 37 °C for 10 min, and RPA amplification at 37 °C for 30 min. The forward primer (FP) and reverse primer (RP) bind with ligation products, and SYTO 9 inserts into the base pair of dsDNA. (**B**) probes of miR-1273, miR-296, and miR-29 are used in ligation-RPA assay. (**C**) Validation of miR-1273 Ligation product using gel image. Ligation products show an intense mark near 75 bp. (**D-F**) Delta Relative fluorescence signal (ΔRFU) for Positive samples for miR-1273, miR-296, and miR-29, respectively. The threshold (Th) was set using µ+3σ= 7997 A.U., where µ and σ indicate the mean and standard deviation, respectively.

To validate the ligation assay, we verified the ligation reaction by testing the miR-1273, P1, P2, and ligation products using agarose gel electrophoresis (**Fig. 1C**). The results showed that the ligation product band was intense and brighter than the miR-1273, P1, and P2 bands because the larger, high-molecular-weight ligation products moved more slowly than the tiny fragments of ssDNA. These results demonstrated that ligation products, including P1 and P2 sequences, were successfully synthesized in the presence of miR-1273. To revalidate the ligation-coupled RPA reaction, we tested 100 pM concentrations of miR-1273, miR-296, and miR-29. We used an interacting fluorescent dye (SYTO 9) in real-time quantitative measurement of RPA assay to determine the change in the Relative Fluorescence Units (ΔRFU) signal of miR-1273, miR-296, and miR-29. **Fig. 1D** showed a noticeable signal time difference between positive controls (PC) and negative controls (NTC) at the threshold lines (µ+3σ= 7997 AU). Ideally, the NTC should not pick up any signal. However, the NTC shows signal lagging from the PC due to non-specific amplification, shown in Supplementary **Fig. S1A-C**. NTC also displayed that the fluorescent signal was increased with the reaction time in the ligation-RPA platform. It indicated that there was a non-specific amplification due to the non-specific binding of probes and primers. Our assay had two probes (P1 and P2) and primers (FP and RP). The P2 can directly hybridize with FP to form double-strand DNA (dsDNA) and non-specific products (**Supplementary Fig. S2A**). Furthermore, P1 and P2 form self/hetero dimers and can be amplified to non-specific products due to cross-dimerization in isothermal amplification (*42–44*)(**Supplementary Fig. S2B**).

To validate our investigation of non-specific bindings from negative signals by self or hetero-dimer amplification, we sequentially added P1, P2, RP, FP, and their various mixtures into the RPA amplification system. Next, a commercial enzyme digestion kit was used to break down proteins, allowing for selective detection of amplicons via gel electrophoresis (**Supplementary Fig. S2C**). It showed that RPA invariably forms large amounts of side products in practice, which come from hybridizing P1 and P2 as well as P2 and FP and probes dimers, called primer-dimers (*44*, *45*). High concentrations of probes encourage off-target interactions because the amplification of probe dimers is more efficient than the amplification of the desired amplicons, and probe–probe interactions eventually eliminate target amplification (*43*, *46*). The time difference between the NTC and PC increased as the reaction progressed due to enhanced molecular interactions between probes and miRNA. In the PC, miRNA-probe interactions delayed dimer formation compared to the NTC. For severe SARS-CoV-2 infection in children, two additional target miRNAs are identified: miR-296 and miR-29. For each potential miRNA, a set of two ligation probes (P1 and P2) were designed for ligation reaction (**Fig. 1B**), and two primers were planned to amplify the ligation products for miR-296 and miR-29 (**Supplementary Table S1, S2, S3**). We also validated the feasibility of the reaction platform for miR-296 and miR-29 for 100 pM concentrations, respectively (**Fig. 1E-F**). The results showed that our platform could detect miR-1273, miR-296, and 29 effectively. **Supplementary Fig. S1B,C** demonstrated the raw signal for miR-296 and 29. Based on the above discussion, we consider using ΔRFU instead of a direct RFU signal for the assay readout. The total detection processes were simple and rapid (<42 min, now: 2 min Hybridization and Annealing, 10 min ligation, and 30 min RPA) without thermal cycles, enabling microRNA detection to apply to SARS-CoV-2 infection in children at POCT.

### Design and Validation of Portbale Analzyer

To translate this assay into a portable analytical tool, we designed and developed a compact, portable fluorescent analyzer and determined the miRNA’s resolution with this device. The portable analyzer consists of 8 detection channels, including 11.5 cm in length, 7.5 cm in width, and 7 cm in height. This device closely resembles our group’s current state-of-the-art model for multiplex target detection (*47–49*). We have re-engineered the device program to support isothermal reactions and single-target detection using the cyan channel (500–520 nm) for single-wavelength measurement, as shown in **Fig. 2A**. The small size of the detection device allows patients to test samples for multiplexed analysis at POCT (*50*, *51*). Our portable device connects to a smartphone via a user-friendly app that enables real-time monitoring of fluorescence curves and result interpretation. As shown in **Fig. 2B**, we integrated a low-cost, compact, and highly sensitive AS7431 color sensor into the device, eliminating the need for conventional optical filters. Specifically, an intercalating dye, SYTO 9, is excited by a blue LED, and the color sensor captures the emission. The device includes an aluminum heating block with an attached MC65F103A NTC 10 kΩ (kilo-Ohm) thermistor and a Raspberry Pi-based control unit that manages the entire process and temperature feedback. Real-time fluorescence intensity is recorded, analyzed, and interpreted through software algorithms, calibrating the results to eliminate baseline offset. The device is powered by a DC jack using a 9V battery. Its filter-free optical design significantly reduces both size and complexity. This miniaturized, user-friendly, and cost-effective device has promising potential for miRNA detection in primary medical facilities.

**Fig. 2.**
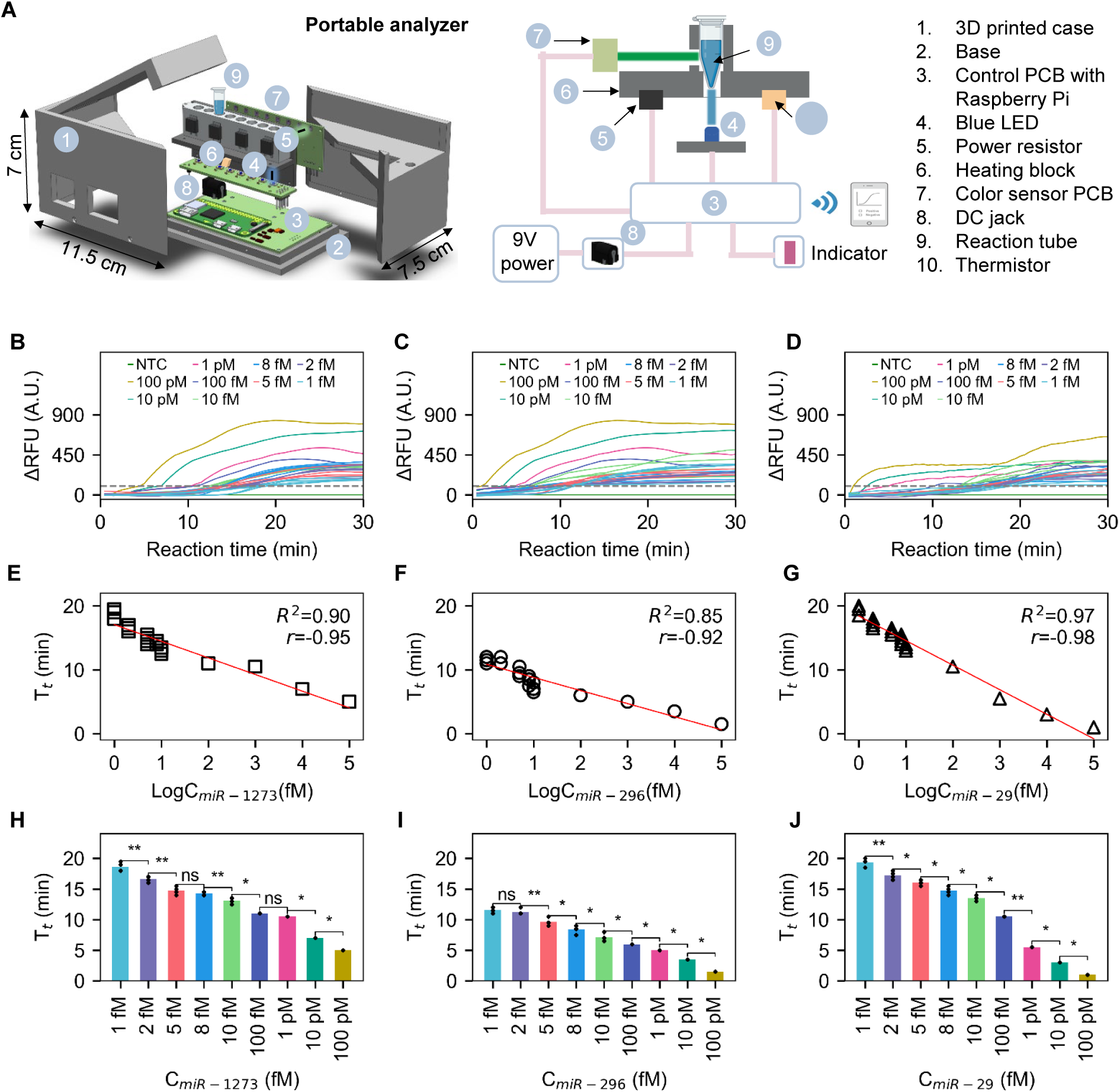
Quantitative performance and sensitivity of the assay. (**A**) Exploded view and schematic of the portable analyzer. (**B-D**) Quantitative analysis showing the ΔRFU of miR-1273, miR-296, and miR-29 at concentrations of 100 pM, 10 pM, 1 pM, 100 fM, 10 fM, 8 fM, 5 fM, 2 fM, and 1 fM, respectively. The gray dashed line represents the threshold (µ + 3σ = 100.43 A.U.), where µ and σ denote the mean and standard deviation. (**E-G**) The assay demonstrates linearity by plotting time-to-positive (Tt) against the logarithm of the concentration for each miRNA target exceeding the threshold. (**H-J**) Assay resolution shown using t-test results to evaluate resolving capabilities. "ns" indicates no significance, while * and ** denote statistically significant differences.

### Analytical Sensitivity in the Portable Device

To evaluate the analytical sensitivity of the assay and resolving capabilities of the portable device, we tested the synthesized miRNA samples, which are miR-1273, 296, and 29 concentrations from 1 fM to 100 pM. For severe SARS-CoV-2 cases, the miRNA concentrations are downregulated and exist at approximately 1 fM (*39*), so resolving 1fM is crucial for the portable analyzer. In **Fig. 2B-D**, following the hybridization/annealing and ligation reaction, we measured the ΔRFU signal of miR-1273, 296, and 29 of the ligation-RPA reaction using the portable analyzer. We tested the concentrations of 100 pM, 10 pM, 1 pM, 100 fM, 10 fM, 8 fM, 5 fM, 2 fM, and 1 fM for all three miRNAs. Four reactions were run for each miRNA for the lower concentrations (10 fM, 8 fM, 5 fM, 2 fM, and 1 fM). We resolved the minimum 1 fM concentrations for each target. We partitioned the miRNAs into three samples and added them into three detection channels, including a negative control in the other channel. Each tube in various channels contained specific probes and primers to ligase and amplify each corresponding miRNA. The sequences of primers are shown in the Supplementary Table **S1-3**. The results showed a noticeable reaction time difference between positive and negative samples for each miRNA over threshold lines (100.43 AU). The threshold was determined using the raw NTC data shown in Supplementary **Fig. S3A-C** and calculating from the µ+3σ, where µ and σ stand for the mean and standard deviation of the signal, respectively. It demonstrated that our principle of ligation-RPA can successfully pick up signals from various miRNAs from 1 fM to 100 pM for multiplexed miRNA detection. In Supplementary **Fig. S4**, the 1 fM signal is closer to the threshold line than the NTC. Below 1 fM, 500 aM and 800 aM did not produce signals above the threshold line.

To validate the quantitative capabilities of the portable device, we evaluate the linearity of the assay. In **Fig. 2E-G**, we calculated time as positive ‘(Tt)’ values when the ΔRFU crosses the threshold line. miR-1273, 296, and 29 displayed R^2^ values of 0.90, 0.85, and 0.97, respectively, and correlation coefficient r-values of 0.95, 0.92, and 0.98, respectively. The R^2^ values indicate a strong proportion of the variance in each measurement. Similarly, r values show a positive correlation of data. These results showed that the ligation-RPA system could effectively detect various concentrations of miRNAs up to 1 fM. This suggests that our portable device is precise, consistent, and reliable for miRNA assay detection. These features make our device well-suited for the portable analysis of multiplex assays. Furthermore, to show the resolution capability of measuring each miRNA target using portable devices, we showed the T-test results with a bar plot. In **Fig. 2H-J**, we found that we can differentiate 1 fM concentration with the T-test result with a p-value <0.05, which means the difference between the two groups is distinguishable at a 95% confidence interval (CI) level. However, the results look similar in two consecutive data points in a few cases. For instance, for miRNA 1273, 5 fM, and 8 fM, and for miRNA 296, 1 fM and 2 fM showed similar kinds of ΔRFU and corresponding T_t_, which labeled "ns" (not significant), showing no statistically significant difference. To clarify, we showed 1 fM to 10 fM results separately in **Supplementary Fig. S3D-F**. The result demonstrated that with the increased concentration of miRNA sample, the T_t_ value decreased, which can be set as a positive reference for the clinical sample to evaluate unknown concentration. It also shows that both the device and assay perform better and are usable for clinically relevant concentrations for severe miRNA.

### Analytical Specificity in the Portable Device

To further validate the cross-reactivity of our assay, the specificity of the platform is essential for detecting specific miRNA in the abundance of human saliva samples. We need to inspect the interferents of the signal of the miRNA assay against the other two types of miRNA targets as inferring targets to investigate the specificity for evaluating the detection capability. High concentrations of miRNAs (10 pM) were used in miRNA-1273, 296, and 29 assays. As illustrated in **Fig. 3A**, the first row displays results for the NTC sample, showing no amplification. In the second row, the specificity test for miR-1273 demonstrates that amplification occurs only with miR-1273 probes (P1 and P2), while no amplification is observed with other probes, such as miR-296 probes and miR-29 probes. Similarly, in the third and fourth rows, specific targets miR-296 and miR-29 only amplify when mixed with their respective probes. In the last row, a triplex sample containing miR-1273, miR-296, and miR-29 produces a signal for all probe types in the ligation-RPA assay, indicating that each target in the triplex sample is detected by its corresponding probe (for example, miR-1273 in the triplex mixture is detected by miR-1273 probes, P1, and P2).

**Fig. 3.**
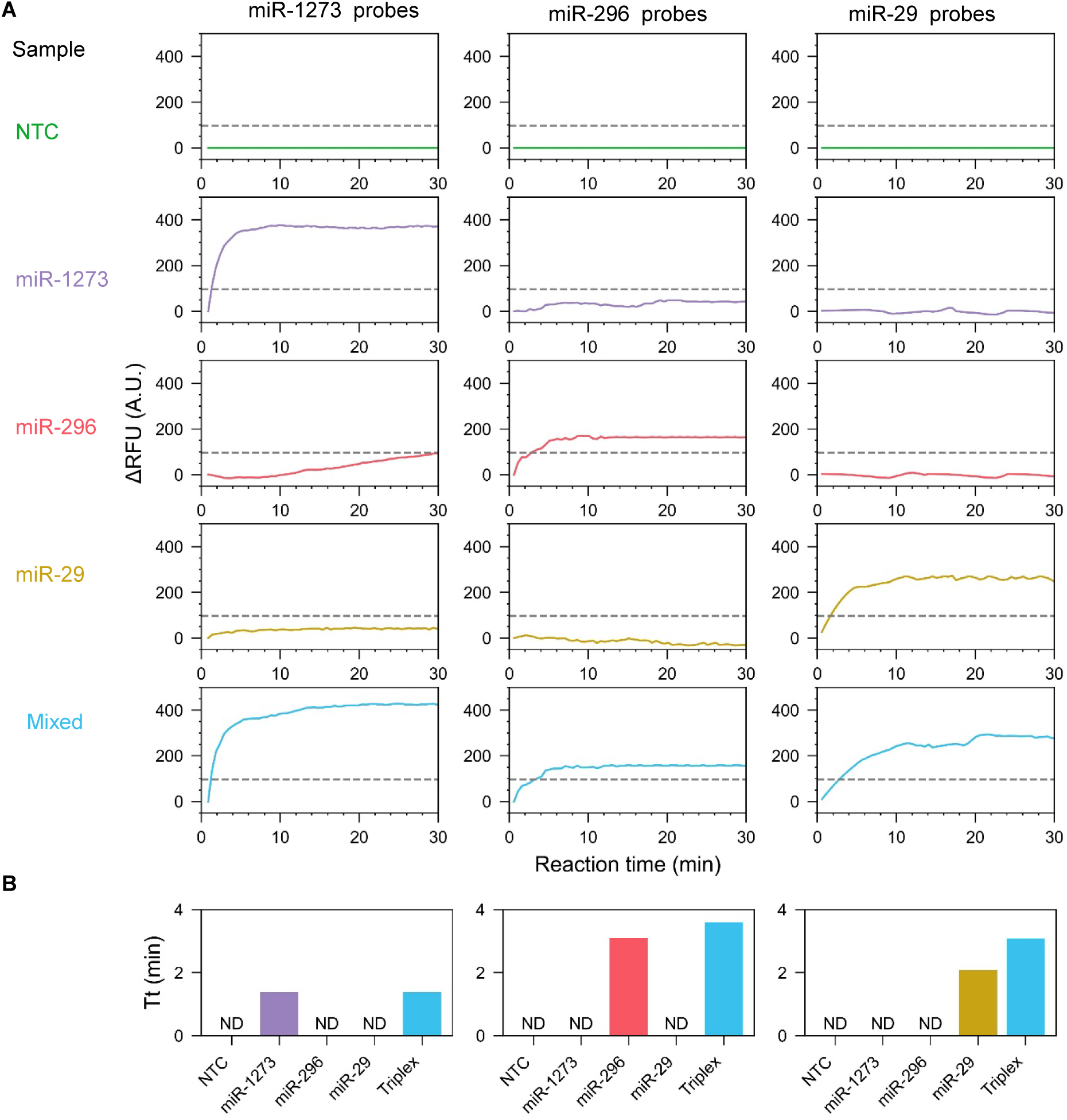
Specificity of the assay. (**A**) ΔRFU signals of NTC, miR-1273, miR-296, miR-29, and mixed miRNAs, represented with different colors. (**B**) Bar plot of Tt values for NTC, miR-1273, miR-296, miR-29, and mixed miRNAs. "ND" indicates "Not Detected," meaning the signals did not cross the threshold (µ + 3σ = 100.43 A.U.). The gray dashed line represents the threshold.

To compare the specificity of each miRNA and the triplex miRNA assay, we present a bar plot of the 𝑇𝑇_𝑡𝑡_ values for five cases: NTC, miR-1273, miR-296, miR-29, and triplex samples. In **Fig. 3B**, the left panel shows that only the miR-1273 and triplex sample signals were detected, with 𝑇𝑇_𝑡𝑡_ values of less than 2 minutes. Similarly, the middle and right panels indicate that signals for miR-296 and triplex and miR-29 and triplex were detected. The reaction time of miR-1273 was approximately similar to the mixture of miRNAs. It indicated that the interfering miRNAs did not affect the miRNA-1273 detection. These results demonstrated that the miR-1273 assay has a high specificity, which could accurately and specifically recognize target miR-1273. Finally, we investigated the effect of miR-296 and 29 on miR-1273. The results exhibited 100% specificity of the assay. These results demonstrated that our platform could analyze clinically complex miRNA samples.

### Design and Validation of Portable miRNA Extraction

To evaluate the performance of our assay in detecting viral presence in clinical and field scenarios, we conducted an extended sensitivity comparison against standard benchtop extraction methods. Using multiplexed miRNA detection on a portable extraction system, we tested eight clinical samples, including two severe and six non-severe cases, with positive reference concentrations of 100 fM, 10 fM, and 1 fM. Saliva swabs were collected with DNA Genotek kits (proprietary nucleus acid solution). Our group developed an integrated platform with extraction reagents and portable centrifuge devices to purify miRNAs in saliva. This semi-automated platform allows simple users to extract miRNAs from saliva (*52*, *53*). We employed the commercial miRNA extraction Kit to extract all the miRNAs from saliva to obtain the practical content of target miRNAs. **Fig. 4A** shows the workflow of the multiplex detection of clinical samples. The process includes saliva collection, sample extraction using a portable or benchtop centrifuge, and multiplex detection using a portable or benchtop analyzer.

**Fig. 4.**
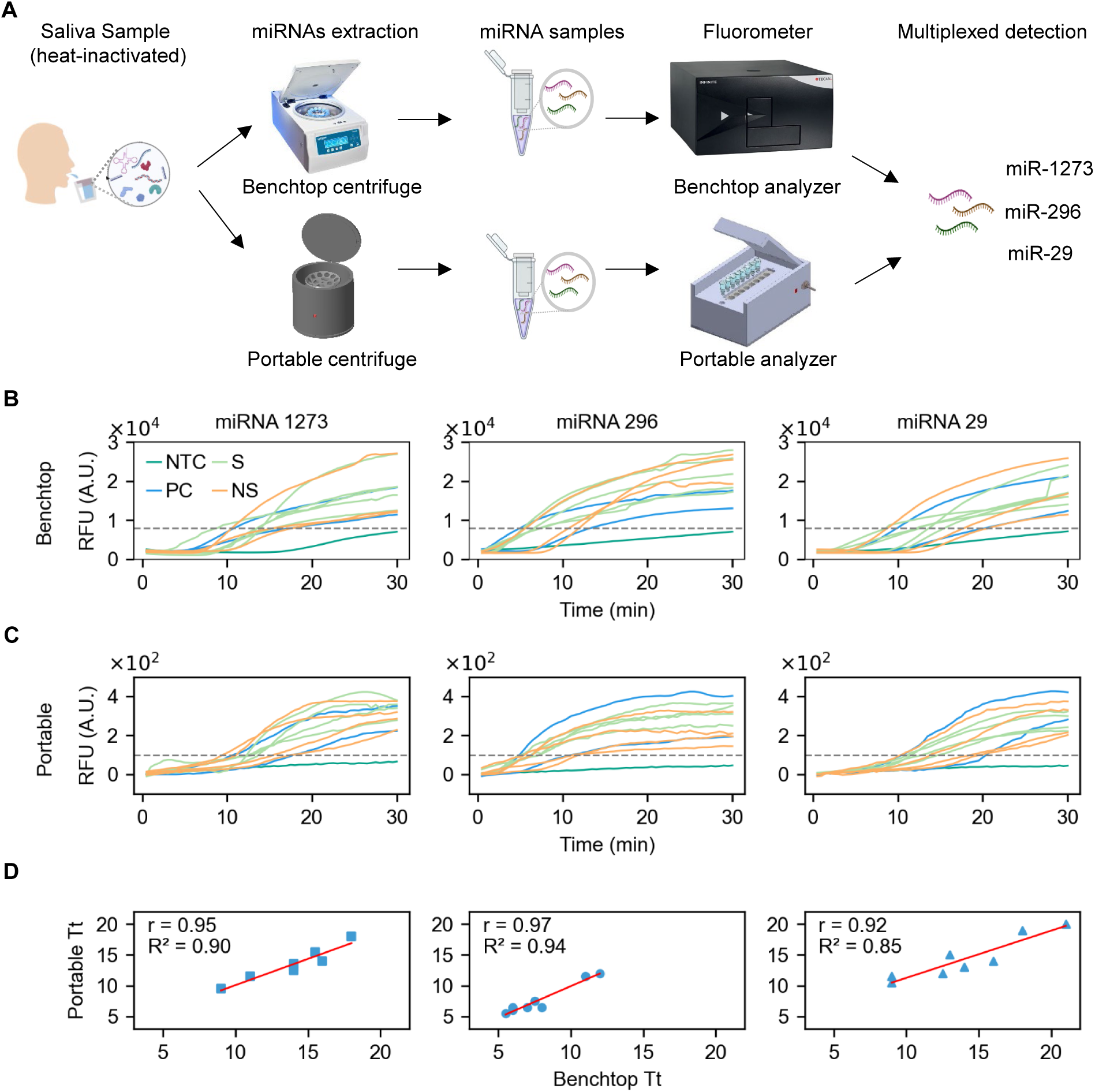
Validation of portable extraction and analyzer. (**A**) Schematic diagram of the workflow, illustrating the process from saliva collection to the readout of multiplex detection. Portable and benchtop extraction and analyzers are used in the intermediate steps. (**B**) RFU values from the benchtop analyzer for six non-severe and two severe samples, with the threshold set at µ + 3σ = 7997 A.U. (**C**) RFU values from the portable analyzer for the same samples, with the threshold calculated as µ + 3σ = 100.43 A.U. (**D**) Comparison of Tt values between the portable and benchtop analyzers, including R² and r values for miR-1273, miR-296, and miR-29, respectively.

First, to compare the performance of our portable analyzer with the traditional Tcan analyzer, we analyzed the amplification curves generated for the miR-1273, miR-296, and miR-29 types on a benchtop centrifuge and analyzer. **Fig. 4B** presents the ΔRFU signal for all eight clinical samples for three different miRNAs using benchtop devices. We parallelly targeted 3 different miRNA samples with 3 PC: 100 fM, 10 fM, 1 fM, and one NTC. The threshold was set to 7997 A.U. for this detection. From this threshold, we evaluated the T_t_ values for all samples. Furthermore, we need to verify the portable centrifuge device’s workability because it is running for the first time for miRNA samples. Each sample was divided into two equal aliquots of 300 µL; one was extracted using a benchtop centrifuge, and a portable centrifuge was used for the other. **Fig. S5** presents the total RNA concentrations of 6 non-severe and 2 severe clinical samples, and the results are compared with those of the benchtop centrifuge. The portable centrifuge shows a strong linear fit with R^2^ values of 0.93 and a strong positive correlation with an r-value of 0.97. After successful extraction, we used our portable analyzer to acquire the RFU signal from the assay.

To demonstrate the quantification capabilities of the portable analyzer for eight clinical samples, we processed the miRNA extracted using the portable centrifuge through the portable analyzer to detect miR-1273, miR-296, and miR-29. **Fig. 4C** presents the ΔRFU signal for all eight clinical samples for three different miRNAs using portable extraction and analyzer. The threshold value was set at 100.43 A.U. Finally, to demonstrate the consistency between the benchtop and portable systems, we compared the T_t_ values presented in **Fig. 4D**. The R² values for miR-1273, miR-296, and miR-29 were 0.90, 0.94, and 0.85, respectively, with corresponding r values of 0.95, 0.97, and 0.92. These results confirm the strong concordance between the two systems, highlighting the portable device’s capability to deliver results comparable to the benchtop system with high accuracy and reliability. Our results establish the applicability of the portable extraction system as an alternative to conventional laboratory methods. The portable analyzer demonstrated exceptional utility for accurately quantifying miRNAs. The multiplex assay’s high sensitivity, specificity, and consistency in a portable format make it an invaluable tool for real-time diagnostics of clinical SARS-CoV-2 miRNA samples, especially in areas where traditional laboratory infrastructure is unavailable.

### Downregulation of miRNAs in Severe Clinical Samples

To find out the profiling and the prediction of unknown severe and non-severe clinical SARS-CoV-2 miRNA, we tested 154 clinical SARS-CoV-2 samples. The detection of miRNA in concentrations within the femtomolar (fM) level requires high sensitivity and accuracy (*54*). We tested 43 severe and 111 non-severe clinical samples with internal PC samples of 100 fM, 20 fM, 10 fM, and 1 fM. **Fig. 5A-C** shows the quantitative measurement of all 154 samples for miR-1273, miR-296, and miR-29. The results demonstrated that all miRNAs underwent successful ligation and RPA amplification, with each crossing the detection threshold.. Next, to determine the concentrations of 154 unknown samples, we utilized PC for calibration. Using the T_t_ values obtained from PC samples with known concentrations of 100 fM, 20 fM, 10 fM, and 1 fM, we calibrated the concentrations of miRNA targets in the 154 unknown severe and non-severe samples. **Fig. 5D-F** presents the calibration results of miR-1273, miR-296, and miR-29, respectively. We measured the R² values for miR-1273, miR-296, and miR-29, which were 0.98, 0.90, and 0.94, respectively, with corresponding r values of 0.99, 0.95, and 0.97. The result indicates that the calibration method is highly accurate and reliable, demonstrating strong linearity and correlation.

**Fig. 5.**
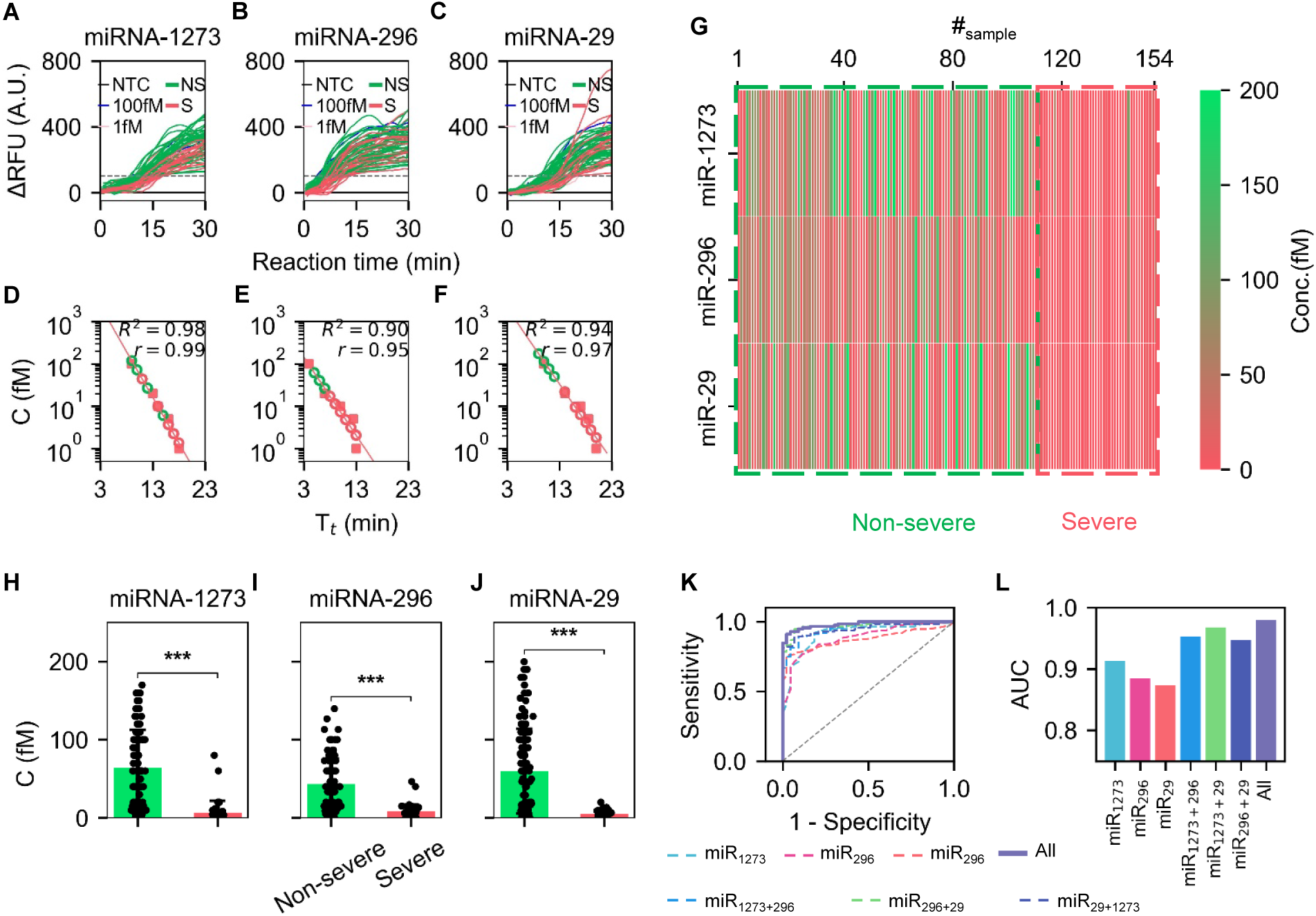
Testing of clinical raw salivary samples. (**A-C**) ΔRFU signals of miR-1273, miR-296, and miR-29 from 154 clinical samples, including 111 non-severe and 43 severe cases. The gray dashed line represents the threshold (µ + 3σ = 100.43 A.U.). Green and red colors indicate non-severity and severity, respectively. (**D-F**) Calibration plots determine unknown Tt values using known Tt values. (**G**) Heat map presenting the 154 clinical samples, with a red-to-green color gradient indicating low to high concentrations. (**H-J**) Bar plots comparing non-severe and severe samples, where *** indicates statistical significance with a p-value < 0.001. (**K**) The ROC-AUC plot shows that combined miRNA testing outperforms individual and dual combinations miRNA testing regarding AUC values. (**L**) Barplot to show the comparison of the single, dual, and combined miRNA testing results.

To visualize the concentration profile of miR-1273, miR-296, and miR-29, a heat map of 154 clinical samples is shown in **Fig. 5G**. From the heat map, we can easily see that the severe concentration is much smaller than that of the non-severe sample. The severe samples are reddish, whereas the non-severe samples are greenish. The severe miRNA concentrations are mostly less than 10 fM, whereas the non-severe sample has many miRNA variations from 10 fM to 200 fM. Notably, the saliva of children with severe illness exhibited significantly decreased miR-1273, miR-296, and miR-29 levels (*3*). The miR-1273, miR-296, and miR-29 are significant early severe SARS-CoV-2 infection markers. The results show that miRNA levels in severe cases are less than in non-severe cases, with most of miR-1273, miR-29, and miR-296 downregulated.

To differentiate severe and non-severe samples, we plotted bar plots and conducted T-test between them for each miRNA sample in **Fig. 5H-J**. The *P* values are 2.34 × 10^-9^, 5.28 × 10^-7^, and 5.54 × 10^-8^, respectively. These results indicate strong evidence against the null hypothesis, confirming statistically significant differences between the groups for all three miRNAs. These results suggest that the observed effects are highly unlikely to be due to random chance. The mean concentrations for miRNA-1273, 296, and 29 are 66.32 fM, 38.72 fM, and 63.85 fM for non-severe samples, while 6.82 fM, 7.47 fM, and 4.75 fM for severe samples, respectively. In children with severe SARS-CoV-2 infection, the levels of salivary miRNA are downgraded compared to those with milder infections, with most miRNAs showing reduced expression.

Finally, to assess the trade-off between the sensitivity and specificity of measurement, we evaluate whether detecting a single miRNA is sufficient or if combining two or three improves assay performance. **Fig. 5K** compares single, dual, and combined miRNA detections, assessing their ROC-AUC plot values across varying thresholds. To evaluate the AUC for single miRNAs, we used the raw concentration values of each miRNA as input features, assigning binary labels (1 for non-severe and 0 for severe) to the samples. We computed the ROC curve by plotting the true positive rate (sensitivity) against the false positive rate (1-specificity) across various thresholds. We calculated the AUC value to quantify each miRNA’s ability to differentiate between the two groups. Additionally, for the dual and combined evaluation of miRNAs, we trained a logistic regression model using the concentrations of miR-1273, miR-296, and miR-29 as input features and sample labels (1 for non-severe, 0 for severe) as outputs. The model assigned weights to each miRNA based on its contribution to the classification and generated predicted probabilities (scores) for the non-severe class. These scores were used to compute the ROC curve and the AUC, reflecting the dual and combined discriminatory power of either two or three miRNAs in distinguishing severe from non-severe samples. The AUC values for the combined-miRNA AUC of 0.98 outperform the two-miRNA combinations (0.95, 0.96, and 0.94) and the individual miRNA AUCs (0.91, 0.88, and 0.87), highlighting the improved accuracy of multi-miRNA detection, shown in **Fig. 5L**. These results indicate that multiplexing enhances specificity and sensitivity compared to single-miRNA testing, albeit at a higher cost. Differentiating between non-severe and severe samples is crucial for accurately assessing the severity of the condition of children’s infection of SARS-CoV-2, which can help in more effective treatment for disease detection in a POC setting.

## Discussion

The outbreaks in SARS-CoV-2 cases among children have highlighted the critical need for rapid, reliable, and highly sensitive nucleic acid-based POC diagnostic tools that can be used either for at-home testing or on-site epidemic monitoring. Saliva, a non-invasive collection medium, shows great potential for such applications. The relative abundance of salivary miRNA shows potential biomarkers, with levels notably downregulated in severe SARS-CoV-2 infections compared to mild cases. However, detecting miRNA linked to severe disease remains challenging due to its low concentration in clinical specimens. Among various isothermal amplification approaches available for miRNA detection, RPA stands out for its ease of use and the reduced number of primers required for amplification. Nevertheless, direct RPA amplification of miRNA has consistently faced limitations because miRNA sizes are between 19 and 24 bases. A two-stage additional step, hybridization/annealing and ligation reaction, enhances the structural integrity of the target by generating a partially double-stranded DNA configuration, enabling more effective RPA amplification. In this work, we have developed an isothermal ligation-RPA assay for multiplexed detection of miRNAs (miR-1273, miR-296, and miR-29) without thermal cycling, using a portable analyzer with a total assay time of 42 minutes: 2 minutes for annealing, 10 minutes for ligation, and 30 minutes for RPA (**Fig. 1A**). Although the RPA assay was conducted for 30 minutes, most of the miRNA assay showed the amplification within 20 minutes. However, this RPA assay might be costly in terms of reagent collection and usage.

The high sensitivity of the assay is essential for miRNA detection, as miRNAs play a critical role in the host’s response to infections by regulating proteins involved in the immune system (*55*). The concentrations of miRNA markers in biofluids can be extremely low, often at the picomolar to femtomolar level, particularly during the early stages of the disease (*40*). Recent work shows that target sites for 28 miRNAs have been identified on the SARS-CoV-2 genome, indicating that the virus may act as a "sponge" to suppress immune-regulating miRNAs and potentially weaken the host’s immune response (*18*). Among these, miR-4495, miR-296-5p, miR-548ao-3p, and miR-1273c showed downregulation from mild to severe infection (*3*). Here, we developed a portable analyzer to detect and resolve the miRNA at femtomolar concentrations (**Fig. 2**), which aligns with the concentration range found in severe saliva samples for SARS-CoV-2 detection in children. The device has eight channels. To detect miRNA, we used three channels for parallel target detection; other channels acted as PC and NC. The portable analyzer used blue light to excite the SYTO 9 dye, and we recorded the color using a color sensor. However, a one-pot assay is always needed to call it proper to save time and reagents. The future scope will be engineering the appropriate probe concentration ratio for one-pot-assay.

Specificity is another essential term that correctly indicates the true negative samples. When using multiple probes and primers, ensuring that none cross-react with non-target sequences is vital, maintaining exclusivity to their specific target. Herein, our assay exhibited 100% specificity for target miRNAs, even in the presence of interfering targets (**Fig. 3**). With this performance, we showed the portable device showed performance identical to that of the benchtop extraction and analyzer. In our comparison of eight samples (four non-severe and four severe), the strong linearity of measurements suggests that both the portable and benchtop extraction methods are highly reliable and unlikely to introduce errors (**Fig. 4**).

Furthermore, we were confident in advancing to clinical sample testing based on the results. A total of 154 clinical samples were analyzed in **Fig. 5**, comprising 111 non-severe and 43 severe cases. Our measurements revealed that miR-1273, miR-296, and miR-29 were significantly downregulated from non-severe to severe cases. The p-value between the two groups (p<0.001) indicates a statistically significant difference, highlighting the robustness and reliability of the results. Notably, the combined detection of the three miRNAs provided greater diagnostic confidence than single- or two-miRNA testing, achieving an AUC value of 0.98 in the combined ROC analysis. This highlights the enhanced sensitivity and specificity of multi-miRNA detection. However, while multiplex detection improves accuracy, it also increases costs, making a trade-off between sensitivity, specificity, and affordability, which is crucial for clinical diagnostics. We envision that this ligation-RPA-based portable analyzer could serve as a versatile platform for simple, rapid, and multiplex miRNA detection, with significant potential for point-of-care (POC) diagnostics and early detection of severe SARS-CoV-2 infections in children, as well as other miRNA-related diseases. While testing three miRNAs may incur higher costs, it offers superior confidence in distinguishing severe miRNA-associated conditions.

Several further improvements are necessary to enhance the applicability of our approach for POC miRNA detection. Current methods rely on liquid-phase reagents, necessitating the development of reagent lyophilization techniques for bedside deployment. The existing workflow involves multiple manual steps, including using a portable centrifuge for semi-automated lysis from saliva samples. After lysis, the extracted miRNA must be transferred using a pipette for subsequent steps. The workflow also requires handling solutions during temperature transitions, such as moving the sample from an 80°C hybridization bath to an ice-cool bath and manually adding the RPA mastermix after ligation in separate steps. To address these challenges, a fully integrated “sample-in, answer-out” microfluidic chip could be designed to automate sample lysis, solution transfer, hybridization, ligation, and RPA reactions. This integration would simplify the workflow, making our portable analyzer a powerful tool for the early detection of SARS-CoV-2 miRNAs in children, particularly in POC settings.

## Methods and Methods

### Materials and Chemicals

miRNA sequences, primers, and probes were synthesized, and nuclease-free water was purchased from Integrated DNA Technologies (IDT). Clinical samples were supplied from Dr. Hicks’s Lab and collected at Central Michigan University and the University of Pittsburgh Medical Center (UPMC). DNA purification kits (#28104) and miRNA extraction kits (#217004) were purchased from Qiagen. The RPA amplification kit, TwistAmp® Liquid Basic (#TALQBAS01), was purchased from TwistDx. Gel Loading Dye, Purple (6X) (#B7025), SplintR ligase (# M0375L), 10x T4 DNA Ligase Reaction Buffer (#B0202S), and extreme thermostable single-stranded DNA binding protein (ET SSB, # M2401S) were both obtained from New England Biolabs Inc (NEB). SYBR™ Safe DNA Gel Stain (#S33102), Ultra Low Range DNA Ladder (#10597012), dNTP mix (10 mM, #R0192), 200 µL sterile pipette tips (#02707430) and SYTO™ 9 Green Fluorescent Nucleic Acid Stain (100mL, # S34854) were bought from Thermo Fisher Scientific (Thermo Scientific, USA). Chloroform (#366927) was purchased from Millipore Sigma.

### Saliva Collection

This prospective cohort study involved a convenience sample of children under 18 years of age evaluated in the emergency departments of two children’s hospitals between March 2021 and February 2022. Children were excluded if they were pregnant, had recent dental infections, head or neck trauma, active seizures, presented for psychiatric evaluation, or were accompanied by a non-English or non-Spanish speaking guardian. Saliva samples were collected using a standardized procedure after an oral rinse with tap water. A highly absorbent swab was placed in the sublingual and parotid regions for 10-20 seconds. The samples were mixed with a nucleic acid stabilization reagent from Genotek kits (Catalog #: ORE-100, Kanata, ON, Canada) to deactivate the virus and preserve RNA integrity. The research team, trained through manufacturer-provided video instructions, ensured correct sample acquisition. Samples were stored at room temperature and shipped monthly to the Core facility for processing. Quality control measures were strictly followed, with samples failing to meet criteria being re-analyzed or excluded from further analysis

### Ligation-RPA-based miRNA Amplification

#### Design

Our designed target, probes, and primers were aligned precisely for target miRNA 1273, 296, and 29 and confirmed the specificity of the sequence by BLAST search. The PrimerQuest Tool of IDT and the Python-based package PrimedRPA (*56*) were used to design primers and probes. All sequences are summarized in **Table S1-3**.

#### Hybridization and Annealing

The Hybridization reaction of the annealing mixture was a total of 8 mL volume, including the final concentration of 100 nM of 4 mL miRNA, 2 mL of probe P1, and 2 mL of probe P2 was heated to 85 °C for 2-3 min and immediately placed on ice for annealing. The detailed recipe is discussed in **Table S4**.

#### Ligation

The ligation reaction, with a total volume of 2 mL, including 0.2 mL 10x ligase buffer,

0.1 mL SpintR ligase enzyme, 0.13 mL ET SSB enzyme, and 1.57 mL water. A ligation reaction was done in 15 min at 37 ^°^C. Finally, 2 mL of ligase products were mixed with 8 mL annealed samples. The detailed recipe is discussed in **Table S4**.

#### RPA assay

The total volume of RPA reaction is 25 mL, where 5 mL ligation products were added to the 20 mL RPA liquid master mix. RPA mixture contained 0.425 mL of 1.5 mM both primers, 12.5 mL 2×reaction buffer, 1.15 mL of 10 mM dNTPs, 2.5 mL 10×Basic E-mix, 1.25 mL 20×core reaction, 0.5 mL of 100-500 mM SYTO 9, and lastly, 1.25 mL 280 mM MgOAc. The mixture with miRNA was then performed on the TCAN plate reader or portable analyzer at 37 °C for 20 min, with fluorescence acquired every 30 s. The detailed recipe is discussed in **Table S4,** and the reaction cost for a single run is provided in **Table S5**.

#### Gel analysis

The nucleic acids were extracted from the ligation products and amplicons from RPA and detected by the 5% agarose gel electrophoresis performed at 5 V/cm using SYBR™ Safe DNA Gel Stain. The required bands are then analyzed using the BIO-RAD GelDoc Go Imaging System with a 20-second exposure.

### Instrumentation

#### Portable analyzer

The analyzer was developed using PTC Creo software and 3D fabricated with a MakerBot Method X printer. The printed circuit boards (PCB) were designed using Autodesk Eagle and produced by OSH Park. We assembled the electronic components and the Microcontroller Unit (MCU) onto the PCB using soldering. The thermal module comprises a resistive-heating element (PWR263S-20-2R00J, Digi-Key) affixed to the underside of the aluminum heating block using a thermal compound (AATA-5G, Arctic Alumina). A thermistor (95C0606, Digi-Key) is used in the core of the heating block for the feedback system. An N-channel power MOSFET (63J7707, Digi-Key) controls the temperature stability using a negative feedback mechanism. The optical module was designed using an adjustable Res Cermet Trimmer (3296W, Digi-Key) for LED intensity control, a broad-spectrum IC Color Sensor (AS7341, Digi-Key) for optical detection, and a blue SMD LED (1497-1138-1, Digi-Key) for fluorescence excitation. A voltage regulator is used for Power stabilization for the optical components, including several capacitors. An I2C multiplexer expanded the sensor’s capacity. The smartphone GUI, created with MIT App Inventor, enables control and communication with the device via Bluetooth. The cost of Materials for the analyzer is provided in **Table S6**.

#### Portable Centrifuge

The centrifuge was designed using PTC Creo and 3D fabricated using a MakerBot Method X printer (*52*). Soldering and jumper wires were used to connect the internal components of the device. The DC motor was purchased from Amazon (Autotoolhome 12V), the microcontroller used was an Arduino Nano (Arduino.cc), and a 28.8 Wh Li-ion battery was purchased from Digikey (Daytona Industries). All other electronic components, resistors, MOSFETs, switches, LEDs, and buttons, were purchased from Digikey..

### Clinical Sample Extraction

Portable extractions were achieved using the miRNeasy Mini Kit from Qiagen. Following their quick-start protocol, extractions were performed with 350 µL sample, 700 μl QIAzol Lysis Reagent, 900 µL 100% Ethanol, 700 μl Buffer RWT, 1000 μl Buffer RPE, 500 µL 95% Ethanol, and 40 µL RNase-free water. During all extraction stages, the portable device runs at max speed (6000 rpm/ 1743 rcf.) (*52*).

### Data analysis method and statistics

All data processing and figure generation were completed using Python and Origin 2021. The threshold was analyzed using a triplicate of NTC samples with μ + 3σ. Positive samples are classified using Time to Positive (Tt) when RFU reaches a threshold of μ + 3σ. Severe and non-severe concentration was calculated with T-test statistics results using where (*p*<0.05). Correlation and linearity were computed using SciPy and least squares regression.

## Data Availability

All data produced in the present study are available upon reasonable request to the authors

## Acknowledgments Funding

This work was partially supported by the National Institutes of Health (R33HD105610, R33AI147419) and the National Science Foundation (2319913). Any opinions, findings, conclusions, or recommendations expressed in this work are those of the authors and do not necessarily reflect the views of the National Science Foundation and National Institutes of Health.

## Author Contributions

ZZ and MAA designed and validated the assay. MAA performed clinical sample testing and validation. MAA, ZZ, and FG conducted data analysis. AK developed the portable analytical device. AJP developed the portable sample extraction device. SH, US, and SS collected clinical samples and provided clinical insight into the miRNA significance related to COVID-19 severity. WG developed the concept and supervised the overall experiment design. MAA, ZZ, and WG co-wrote the manuscript with input from others.

## Data and materials availability

All data needed to evaluate the conclusions in the paper are present in the paper and/or the Supplementary Materials.

## References

1. C. Huang, Y. Wang, X. Li, L. Ren, J. Zhao, Y. Hu, L. Zhang, G. Fan, J. Xu, X. Gu, Z. Cheng, T. Yu, J. Xia, Y. Wei, W. Wu, X. Xie, W. Yin, H. Li, M. Liu, Y. Xiao, H. Gao, L. Guo, J. Xie, G. Wang, R. Jiang, Z. Gao, Q. Jin, J. Wang, B. Cao, Clinical features of patients infected with 2019 novel coronavirus in Wuhan, China. The Lancet 395, 497–506 (2020).

2. B. Hu, H. Guo, P. Zhou, Z.-L. Shi, Characteristics of SARS-CoV-2 and COVID-19. Nat. Rev. Microbiol. 19, 141–154 (2021).

3. S. D. Hicks, D. Zhu, R. Sullivan, N. Kannikeswaran, K. Meert, W. Chen, S. Suresh, U. Sethuraman, Saliva microRNA profile in children with and without severe SARS-CoV-2 Infection. Int. J. Mol. Sci. 24, 8175 (2023).

4. S. M. Murray, A. M. Ansari, J. Frater, P. Klenerman, S. Dunachie, E. Barnes, A. Ogbe, The impact of pre-existing cross-reactive immunity on SARS-CoV-2 infection and vaccine responses. Nat. Rev. Immunol. 23, 304–316 (2023).

5. CDC, Child mortality and COVID-19, United Nations Children’s Fund (2023). https://data.unicef.org/topic/child-survival/covid-19/.

6. H. Huang, C. Fan, M. Li, H.-L. Nie, F.-B. Wang, H. Wang, R. Wang, J. Xia, X. Zheng, X. Zuo, J. Huang, COVID-19: A Call for Physical Scientists and Engineers. ACS Nano 14, 3747–3754 (2020).

7. N. L. Welch, M. Zhu, C. Hua, J. Weller, M. E. Mirhashemi, T. G. Nguyen, S. Mantena, M. R. Bauer, B. M. Shaw, C. M. Ackerman, S. G. Thakku, M. W. Tse, J. Kehe, M.-M. Uwera, J. S. Eversley, D. A. Bielwaski, G. McGrath, J. Braidt, J. Johnson, F. Cerrato, G. K. Moreno, L. A. Krasilnikova, B. A. Petros, G. L. Gionet, E. King, R. C. Huard, S. K. Jalbert, M. L. Cleary, N. A. Fitzgerald, S. B. Gabriel, G. R. Gallagher, S. C. Smole, L. C. Madoff, C. M. Brown, M. W. Keller, M. M. Wilson, M. K. Kirby, J. R. Barnes, D. J. Park, K. J. Siddle, C. T. Happi, D. T. Hung, M. Springer, B. L. MacInnis, J. E. Lemieux, E. Rosenberg, J. A. Branda, P. C. Blainey, P. C. Sabeti, C. Myhrvold, Multiplexed CRISPR-based microfluidic platform for clinical testing of respiratory viruses and identification of SARS-CoV-2 variants. Nat. Med. 28, 1083–1094 (2022).

8. M. A. Ahamed, A. J. Politza, T. Liu, M. A. U. Khalid, H. Zhang, W. Guan, CRISPR-based strategies for sample-to-answer monkeypox detection: current status and emerging opportunities. Nanotechnology 36, 042001 (2025).

9. Z. Zhang, T. Liu, M. Dong, Md. A. Ahamed, W. Guan, Sample-to-answer salivary miRNA testing: New frontiers in point-of-care diagnostic technologies. WIREs Nanomedicine Nanobiotechnology 16, e1969 (2024).

10. D. Najjar, J. Rainbow, S. Sharma Timilsina, P. Jolly, H. De Puig, M. Yafia, N. Durr, H. Sallum, G. Alter, J. Z. Li, X. G. Yu, D. R. Walt, J. A. Paradiso, P. Estrela, J. J. Collins, D. E. Ingber, A lab-on-a-chip for the concurrent electrochemical detection of SARS-CoV-2 RNA and anti-SARS-CoV-2 antibodies in saliva and plasma. *Nat*. Biomed. Eng. 6, 968–978 (2022).

11. M. Song, H. Bai, P. Zhang, X. Zhou, B. Ying, Promising applications of human-derived saliva biomarker testing in clinical diagnostics. Int. J. Oral Sci. 15, 2 (2023).

12. J. Hindson, Salivary miRNA signature for ESCC. Nat. Rev. Gastroenterol. Hepatol. 20, 559–559 (2023).

13. L. Tribolet, E. Kerr, C. Cowled, A. G. D. Bean, C. R. Stewart, M. Dearnley, R. J. Farr, MicroRNA Biomarkers for Infectious Diseases: From Basic Research to Biosensing. Front. Microbiol. 11, 1197 (2020).

14. R. Ojha, R. Nandani, R. K. Pandey, A. Mishra, V. K. Prajapati, Emerging role of circulating microRNA in the diagnosis of human infectious diseases. J. Cell. Physiol. 234, 1030–1043 (2019).

15. Z. Zhang, Md. A. Ahamed, D. Yang, Biological properties and DNA nanomaterial biosensors of exosomal miRNAs in disease diagnosis. Sens. Diagn., 10.1039.D4SD00373J (2025).

16. R. Garcia-Martin, G. Wang, B. B. Brandão, T. M. Zanotto, S. Shah, S. Kumar Patel, B. Schilling, C. R. Kahn, MicroRNA sequence codes for small extracellular vesicle release and cellular retention. Nature 601, 446–451 (2022).

17. T. Abu-Izneid, N. AlHajri, A. M. Ibrahim, Md. N. Javed, K. M. Salem, F. H. Pottoo, M. A. Kamal, Micro-RNAs in the regulation of immune response against SARS CoV-2 and other viral infections. J. Adv. Res. 30, 133–145 (2021).

18. R. Bartoszewski, M. Dabrowski, B. Jakiela, S. Matalon, K. S. Harrod, M. Sanak, J. F. Collawn, SARS-CoV-2 may regulate cellular responses through depletion of specific host miRNAs. Am. J. Physiol.-Lung Cell. Mol. Physiol. 319, L444–L455 (2020).

19. H. Kobayashi, R. H. Singer, Single-molecule imaging of microRNA-mediated gene silencing in cells. Nat. Commun. 13, 1435 (2022).

20. M. Bentahir, J. Ambroise, C. Delcorps, P. Pilo, J. L. Gala, Sensitive and Specific Recombinase Polymerase Amplification Assays for Fast Screening, Detection, and Identification of Bacillus anthracis in a Field Setting. Appl Env. Microbiol 84 (2018).

21. S. Husale, H. H. J. Persson, O. Sahin, DNA nanomechanics allows direct digital detection of complementary DNA and microRNA targets. Nature 462, 1075–1078 (2009).

22. A. Valoczi, Sensitive and specific detection of microRNAs by northern blot analysis using LNA-modified oligonucleotide probes. Nucleic Acids Res. 32, e175–e175 (2004).

23. J. Chen, J. Lozach, E. W. Garcia, B. Barnes, S. Luo, I. Mikoulitch, L. Zhou, G. Schroth, J.-B. Fan, Highly sensitive and specific microRNA expression profiling using BeadArray technology. Nucleic Acids Res. 36, e87–e87 (2008).

24. X. Pan, A. K. Murashov, E. J. Stellwag, B. Zhang, “Monitoring MicroRNA Expression During Embryonic Stem-Cell Differentiation Using Quantitative Real-Time PCR (qRT-PCR)” in RNAi and microRNA-Mediated Gene Regulation in Stem Cells, B. Zhang, E. J. Stellwag, Eds. (Humana Press, Totowa, NJ, 2010; http://link.springer.com/10.1007/978-1-60761-769-3_16)vol. 650 of Methods in Molecular Biology, pp. 213–224.

25. V. Trevino, F. Falciani, H. A. Barrera-Saldaña, DNA Microarrays: a Powerful Genomic Tool for Biomedical and Clinical Research. Mol. Med. 13, 527–541 (2007).

26. A. B. Villaseñor-Altamirano, Y. I. Balderas-Martínez, A. Medina-Rivera, “Review of gene expression using microarray and RNA-seq” in Rigor and Reproducibility in Genetics and Genomics (Elsevier, 2024; https://linkinghub.elsevier.com/retrieve/pii/B9780128172186000085), pp. 159–187.

27. R. Gonzalo, A. Sánchez, “Introduction to Microarrays Technology and Data Analysis” in Comprehensive Analytical Chemistry (Elsevier, 2018; https://linkinghub.elsevier.com/retrieve/pii/S0166526X18300734)vol. 82, pp. 37–69.

28. M. A. U. Khalid, Md. A. Ahamed, M. Dong, A. Kshirsagar, W. Guan, Hydrogel interfaced glass nanopore for high-resolution sizing of short DNA fragments. Biosens. Bioelectron. 268, 116895 (2025).

29. L. Lan, J. Huang, M. Liu, Y. Yin, C. Wei, Q. Cai, X. Meng, Polymerization and isomerization cyclic amplification for nucleic acid detection with attomolar sensitivity. Chem. Sci. 12, 4509–4518 (2021).

30. S. Feng, H. Chen, Z. Hu, T. Wu, Z. Liu, Ultrasensitive detection of miRNA via CRISPR/Cas12a coupled with strand displacement amplification reaction. ACS Appl. Mater. Interfaces 15, 28933–28940 (2023).

31. D. Li, T. Zhang, F. Yang, R. Yuan, Y. Xiang, Efficient and Exponential Rolling Circle Amplification Molecular Network Leads to Ultrasensitive and Label-Free Detection of MicroRNA. Anal. Chem. 92, 2074–2079 (2020).

32. K. M. Koo, E. J. H. Wee, M. Trau, High-speed biosensing strategy for non-invasive profiling of multiple cancer fusion genes in urine. Biosens. Bioelectron. 89, 715–720 (2017).

33. J. Li, J. Macdonald, Advances in isothermal amplification: novel strategies inspired by biological processes. Biosens. Bioelectron. 64, 196–211 (2015).

34. Md. A. Ahamed, M. A. U. Khalid, M. Dong, A. J. Politza, Z. Zhang, A. Kshirsagar, T. Liu, W. Guan, Sensitive and specific CRISPR-Cas12a assisted nanopore with RPA for Monkeypox detection. Biosens. Bioelectron. 246, 115866 (2024).

35. Md. A. Ahamed, W. Guan, CRISPR-assisted solid-state nanopore sensor for rapid and sensitive point-of-care amendable of monkeypox virus detection via RPA amplification. Biophys. J. 123, 145a (2024).

36. B. B. Oliveira, B. Veigas, P. V. Baptista, Isothermal Amplification of Nucleic Acids: The Race for the Next “Gold Standard.” Front. Sens. 2, 752600 (2021).

37. P. G. Preena, T. V. A. Kumar, T. K. Johny, A. Dharmaratnam, T. R. Swaminathan, Quick hassle-free detection of cyprinid herpesvirus 2 (CyHV-2) in goldfish using recombinase polymerase amplification-lateral flow dipstick (RPA-LFD) assay. Aquac. Int. 30, 1211– 1220 (2022).

38. E. J. H. Wee, M. Trau, Simple isothermal strategy for multiplexed, rapid, sensitive, and accurate miRNA detection. ACS Sens. 1, 670–675 (2016).

39. N. Kumar, N. P. Shetti, S. Jagannath, T. M. Aminabhavi, Electrochemical sensors for the detection of SARS-CoV-2 virus. Chem. Eng. J. 430, 132966 (2022).

40. H. Yan, Y. Wen, Z. Tian, N. Hart, S. Han, S. J. Hughes, Y. Zeng, A one-pot isothermal Cas12-based assay for the sensitive detection of microRNAs. *Nat*. Biomed. Eng. 7, 1583– 1601 (2023).

41. N. V. Bhagavan, C.-E. Ha, “Structure and Properties of DNA” in Essentials of Medical Biochemistry (Elsevier, 2015; https://linkinghub.elsevier.com/retrieve/pii/B978012416687500021X), pp. 381–400.

42. A. V. Ivanov, I. V. Safenkova, A. V. Zherdev, B. B. Dzantiev, Multiplex assay of viruses integrating recombinase polymerase amplification, barcode—anti-barcode pairs, blocking anti-primers, and lateral flow assay. Anal. Chem. 93, 13641–13650 (2021).

43. R. J. Meagher, A. Priye, Y. K. Light, C. Huang, E. Wang, Impact of primer dimers and self-amplifying hairpins on reverse transcription loop-mediated isothermal amplification detection of viral RNA. The Analyst 143, 1924–1933 (2018).

44. N. Sharma, S. Hoshika, D. Hutter, K. M. Bradley, S. A. Benner, Recombinase-Based Isothermal Amplification of Nucleic Acids with Self-Avoiding Molecular Recognition Systems (SAMRS). ChemBioChem 15, 2268–2274 (2014).

45. O. Piepenburg, C. H. Williams, D. L. Stemple, N. A. Armes, DNA Detection Using Recombination Proteins. PLoS Biol. 4, e204 (2006).

46. T. Nolan, R. E. Hands, S. A. Bustin, Quantification of mRNA using real-time RT-PCR. Nat. Protoc. 1, 1559–1582 (2006).

47. T. Liu, A. J. Politza, A. Kshirsagar, Y. Zhu, W. Guan, Compact point-of-care device for self-administered HIV viral load tests from whole blood. ACS Sens., acssensors.3c01819 (2023).

48. T. Liu, A. J. Politza, Md. A. Ahamed, A. Kshirsagar, Y. Zhu, W. Guan, Compact Multiplex PCR Device for HIV-1 and HIV-2 Viral Load Determination from Finger-Prick Whole Blood in Resource-Limited Settings. Biosens. Bioelectron., 116997 (2024).

49. A. Kshirsagar, A. J. Politza, W. Guan, Deep Learning Enabled Universal Multiplexed Fluorescence Detection for Point-of-Care Applications. ACS Sens., doi: 10.1021/acssensors.4c00860 (2024).

50. P. B. Luppa, C. Müller, A. Schlichtiger, H. Schlebusch, Point-of-care testing (POCT): Current techniques and future perspectives. TrAC Trends Anal. Chem. 30, 887–898 (2011).

51. Q. Song, X. Sun, Z. Dai, Y. Gao, X. Gong, B. Zhou, J. Wu, W. Wen, Point-of-care testing detection methods for COVID-19. Lab. Chip 21, 1634–1660 (2021).

52. A. J. Politza, T. Liu, A. Kshirsagar, M. Dong, M. A. Ahamed, W. Guan, A Portable Centrifuge for Universal Nucleic Acid Extraction at the Point-of-Care. 4781228 [Preprint] (2024). 10.2139/ssrn.4781228.

53. A. J. Politza, T. Liu, A. Kshirsagar, M. Dong, Md. Ahasan Ahamed, W. Guan, A Portable Device for Lab-Free, Versatile Nucleic Acid Extraction - Protocol. [Preprint] (2024). 10.17504/protocols.io.kxygxyj4wl8j/v1.

54. T. Jet, G. Gines, Y. Rondelez, V. Taly, Advances in multiplexed techniques for the detection and quantification of microRNAs. Chem. Soc. Rev. 50, 4141–4161 (2021).

55. R. E. Drury, D. O’Connor, A. J. Pollard, The clinical application of microRNAs in infectious disease. Front. Immunol. 8, 1182 (2017).

56. M. Higgins, M. Ravenhall, D. Ward, J. Phelan, A. Ibrahim, M. S. Forrest, T. G. Clark, S. Campino, PrimedRPA: primer design for recombinase polymerase amplification assays. Bioinformatics 35, 682–684 (2019).

